# A vulnerability-based approach to human-mobility reduction for countering COVID-19 transmission in London while considering local air quality

**DOI:** 10.1101/2020.04.13.20060798

**Authors:** Manu Sasidharan, Ajit Singh, Mehran Eskandari Torbaghan, Ajith Kumar Parlikad

## Abstract

An ecologic analysis was conducted to explore the correlation between air pollution, and COVID-19 cases and fatality rates in London. The analysis demonstrated a strong correlation (R^2^>0.7) between increment in air pollution and an increase in the risk of COVID-19 transmission within London boroughs. Particularly, strong correlations (R^2^>0.72) between the risk of COVID-19 fatality and NO_2_ and PM_2.5_ pollution concentrations were also found. Although this study assumed the same level of air pollution across a particular London borough, it demonstrates the possibility to employ air pollution as an indicator to rapidly identify the city’s vulnerable regions. Such an approach can inform the decisions to suspend or reduce the operation of different public transport modes within a city. The methodology and learnings from the study can thus aid in public transport’s response to COVID-19 outbreak by adopting different levels of human-mobility reduction strategies based on the vulnerability of a given region.

**One Sentence Summary:** This study introduces air pollution levels as an indicator for a region’s vulnerability to COVID-19 and suggests human-mobility reduction measures.

## Introduction

The current outbreak of novel coronavirus COVID-19 or severe acute respiratory syndrome coronavirus 2 (SARS-CoV-2), has resulted in the World Health Organization (WHO) declaring it as a global pandemic [*1*]. Reported first within the city of Wuhan, Hubei Province of China in December 2019, the COVID-19 exhibits high human-to-human transmissibility and has spread rapidly across the world [*2*]. The human-to-human transmission of COVID-19 can occur from individuals in the incubation stage or showing symptoms, and also from asymptomatic individuals who remain contagious [*3*]. The COVID-19 has been reported to transmit via the inhalation of exhaled respiratory droplets [*4*] that remain airborne for up to 3 hours [*5*]. The long-term exposure to air pollution has been reported to increase the risk of experiencing severe COVID-19 outcomes [*6*]. The extent to which COVID-19 induces respiratory stress in infected individuals may also be influenced by underlying respiratory conditions [*7*] like acute respiratory inflammation, asthma and cardiorespiratory diseases [*8*]. The simultaneous exposure to air pollutants such as particulate matter (PM) and Nitrogen dioxide (NO_2_) alongside COVID-19 virus is expected to exacerbate the level of COVID-19 infection and risk of fatality [*9, 10*]. Moreover, the adsorption of the COVID-19 virus on PM could also contribute to long-range transmission of the virus [*4*]. For example, the 2003 severe acute respiratory syndrome coronavirus 1 (SARS-CoV-1) infected patients who lived in moderate air pollution levels were reported to be 84% more likely to die than those in regions with lower air pollution [*11*]. The aerosol and surface stability of the COVID-19 or SARS-CoV-2 is reported to be similar to that of SARS-CoV-1 [*5*].

Given the limited understanding of the epidemiology of COVID-19, social-distancing and human-mobility reduction measures can contribute greatly to tailoring public health interventions [*12*]. Consequently, countries across the world have enforced lockdowns and other coordinated efforts to reduce human-mobility [*13, 14, 15, 16*]. The UK’s national framework for responding to a pandemic states that public transport should continue to operate normally during a pandemic, but users should adopt good hygiene measures, and stagger journeys where possible [*17*]. Within the UK, London has recorded the highest COVID-19 related fatalities (i.e. 30.2% of UK’s deaths as of 31 March 2020) [*18*]. On 18 March 2020, further to the UK government’s advice, Transport for London (TfL) closed 40 out of 270 London Underground (LU) stations that do not serve as interchanges with other lines and announced a reduced service across its network [*19*]. This is also because 30% of TfL’s drivers, station staff, controllers and maintenance teams were not able to come to work, including those self-isolating or ill with COVID-19 [*20*]. The UK’s current human-mobility reduction response reflects the need to maintain business continuity, near normal functioning of society and enable critical workers to make essential journeys [*17, 21*]. However, a statistically significant association exists between human-mobility through public transport and transmissions of acute respiratory infections (ARI) [*21, 22*]. It was found that using public transport in the UK during a pandemic outbreak has an approximately six-fold increased risk of contracting an ARI [*21*]. Moreover, boroughs with access to LU interchange stations have historically higher pandemic case rates [*22*], as users are exposed to higher number of individuals in comparison to through stations. The methodology and results from this study can be employed to rapidly identify local authorities/regions that are highly vulnerable to COVID-19 and accordingly inform human-mobility reduction measures across the city’s public transport network.

## Results

As the COVID-19 is an evolving pandemic, the available data at the time of writing this paper on 31 March 2020 on COVID-19 morbidity and mortality for different boroughs in London was collected from [*23, 18*]. The fatality rate across each London boroughs was estimated by dividing the number of reported deaths by the number of reported positive COVID-19 cases. The average air pollution data associated with NO_2_ and PM_2.5_ concentration was collected for London boroughs from [*24*]. NO_2_ data was available for 15 boroughs namely Barking and Dagenham, Bexley, Wandsworth, City of London, Croydon, Greenwich, Havering, Hillingdon, Kensington and Chelsea, Lewisham, Reading, Redbridge, Sutton, Tower Hamlets and Westminster. While, the PM_2.5_ data was available only for 8 boroughs (Barking and Dagenham, Wandsworth, City of London, Croydon, Hillingdon, Kensington and Chelsea, Lewisham). An ecologic analysis was conducted to explore the correlation between short-term air pollution and COVID-19 cases and fatality rates. A linear regression model was fitted to the data for the boroughs with more than 100 reported cases and 10 deaths related to COVID-19 as of 31 March 2020. A strong correlation between short-term NO_2_ and PM_2.5_ pollution concentrations and COVID-19 cases with R^2^ values of 0.82 (*COVID-19 cases = −29.345 + 10.306*NO_2_ concentration*) and 0.72 (*COVID-19 cases = −215.63 + 40.997*PM_2.5_ level*) were observed respectively (see Figure 1). In particular, COVID-19 fatality rate increased with increase in short-term air pollution, where a significant correlation between COVID-19 fatality and NO_2_ and PM_2.5_ pollution concentrations with R^2^ of 0.90 (*fatality rate = 1*.*864+ 0*.*5787*NO_2_ level*) and 0.67 (*fatality rate = −7.733+ 2.3399*PM_2.5_ level*) were found (see Figure 2).

**Fig. 1.**
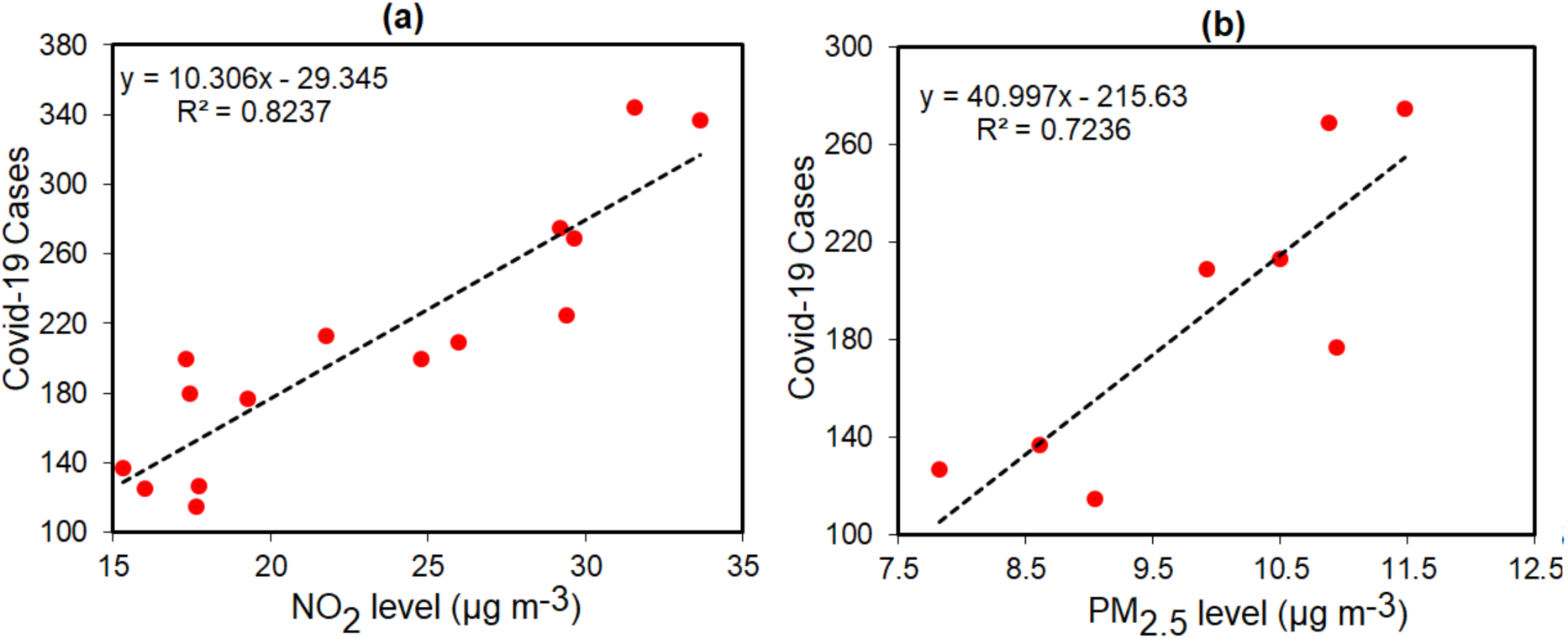
Relationship between **a)** NO_2_ and **b)** PM_2.5_ pollution concentrations and reported COVID-19 cases at London boroughs using data during March 2020.

**Fig. 2.**
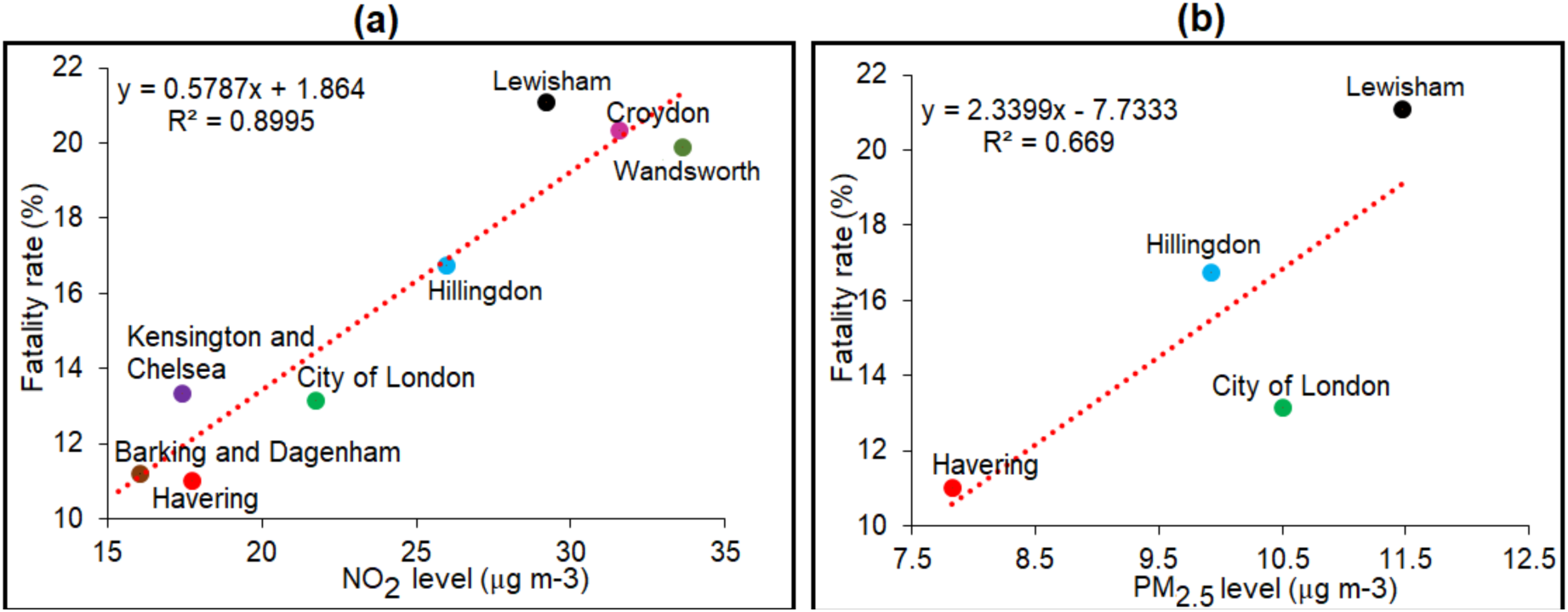
Relationship between **a)** NO_2_ and **b)** PM_2.5_ pollution concentrations and COVID-19 fatality rate for each London borough. The fatality rate was calculating by dividing the number of reported deaths by the number of reported positive COVID-19 cases (associated data was available only for the London boroughs plotted in the figures).

As per TfL’s guidance to LU users [*25*], 40 LU stations across its network were closed as part of the UK Government’s response to COVID-19. The median PM_2.5_ levels recorded by [*26*] for 27 of these 40 closed stations range from 0-50 μg m^−3^ (5 stations), 50-100 μg m^−3^ (9 stations), 100-200 μg m^−3^ (5 stations), 200-300 μg m^−3^(6 stations) and greater than 300 μg m^−3^ (2 stations) (see Table S1). Of the 230 operating stations, the median PM_2.5_ levels recorded for 219 stations range from 0-50 μg m^−3^ (56 stations), 50-100 μg m^−3^ (15 stations), 100-200 μg m^−3^ (15 stations), 200-300 μg m^−3^ (18 stations) and greater than 300 μg m^−3^ (7 stations) [26] (see Table S1). This suggests that approximately 40% of the stations in operation during the current COVID-19 outbreak in London are up to 26 times more polluted than the ambient background locations and the roadside environment (that have median PM_2.5_ level of 14 μg m^−3^ [26]). Moreover, the average NO_2_ concentrations within the LU network was reported to be 51 μg m^−3^ [*27*]; which is 27.5% higher than the NO_2_ limit values for the protection of human health [*28*].

## Discussion

Various studies have reported an association between air pollution levels and excess morbidity and mortality from respiratory diseases [*29, 30*]. Both long-term and short-term exposure to air pollution has been associated with a variety of respiratory conditions [*11*] with children and elderly people being at most risk [*31*]. Our analysis shows that short-term exposure to air pollution (both NO_2_ and PM_2.5_) is significantly correlated with an increased risk of contracting and dying from COVID-19. It was reported by [*6*] that long-term exposure to fine particulate matter (PM_2.5_) increases the risk of COVID-19 deaths. Biologically, either long-term or short-term exposure to air pollutants such as PM_2.5_ and NO_2_ can compromise lung function and therefore increasing the risk of death from COVID-19 [*7*]. Also, the median level of airborne PM_2.5_ in LU during summer is often several times higher than other transport environments such as cycling (35 μg m^−3^), bus (30.9 μg m^−3^), cars (23.7 μg m^−3^) [*32, 26*]. The greatest PM_2.5_ concentrations across the LU network was reported to be on the Victoria Line (around 16 times higher than roadside environment), followed by Northern, Bakerloo and Piccadily line [*26*]. The routine cleaning and maintenance of LU ranges from litter removal to preventing safety incidents rather than on reducing PM concentrations. Our study is in agreement with [*22*] who reported that there are higher pandemic case rates for London boroughs with access to interchange stations, as individuals would interact with more people in comparison to through stations.

An analysis by [*33*] of the UK’s National Health Service (NHS) records have reported that 20% of England’s population are at risk of mortality from COVID-19 due to underlying conditions and age. We support the UK government’s existing COVID-19 guidance [*34*] to exercise good hygiene and to avoid unnecessary travel. While considering the evidence that COVID-19 can be transmitted from an asymptomatic individual [*3*], we do not support the current countermeasure of suspending LU operations on the stations that do not serve as interchanges. This is because of (i) the statistically significant risk of contracting ARI’s on UK’s public transport and higher pandemic case rates within London boroughs that have comparatively easier access to interchange stations [21, 22] and, (ii) relatively higher air pollution levels in LU stations than ambient background locations or road side environment. Since isolating towns or even cities is not yet part of the UK government’s action plan [34], we recommend a vulnerability-based assessment of different boroughs in London and to accordingly suspend or reduce operations on LU stations within highly vulnerable regions. For instance, London Borough of Kensington and Chelsea is seen to be highly vulnerable to COVID-19 fatality from our analysis (see Figure 2a). Table S1 shows that, all the through stations and 3 out of 4 interchange stations (South Kensington, Sloane square, Earl’s court, Notting Hill gate) in this borough are currently operational. Such a vulnerability-based assessment might aid decision-makers in selecting appropriate human-mobility reduction measures to COVID-19 in London’s different local authorities/boroughs (such as apportion of transport staff across railway stations, arranging dedicated shuttling services for key workers, scheduling bus operations etc.) while adhering to the UK’s national framework for response to pandemic outbreaks [*17*].

Given that the immunity to the 2003 SARS-CoV-1 was reported to be relatively short-lived (around 2 years) [*35*], achieving herd immunity for such diseases would be unlikely without overwhelming the healthcare system [*16*]. Moving forward, human-mobility reduction measures provides the greatest benefit to COVID-19 mitigation [*15, 14*] as prevention is potentially cost-effective than cure [*22*] or death. One of the most controversial debate in pandemic countermeasures is the potential benefit of human-mobility reduction and social-distancing attained by closure of public transport systems [*21*]. From a public policy perspective, there is a need to achieve a trade-off between the potential public health benefits of closing public transport during a pandemic thereby delaying the community spread, against the socio-economic impacts of curtailing/reducing human mobility. Determining the vulnerability of regions/locations to COVID-19 might be helpful in achieving such trade-offs. To this end, this study demonstrates that the air pollution levels can serve as one of the indicators to assess a region’s vulnerability to COVID-19. It has to be noted that the number of positive COVID-19 cases considered within this study are only those reported at the hospitals and does not include the growing number of people who are self-isolating due to COVID-19. While the individual risk of contracting and dying from COVID-19 is dependent on various factors (including age, underlying conditions, availability of health care, population density etc.), these results are informative for both scientists and decision-makers in their efforts to reduce the transmission and impact of the ongoing COVID-19 outbreak through appropriate human-mobility reduction strategies.

## Data Availability

All the data used for this study are publicly available.

http://www.londonair.org.uk/london/asp/datadownload.asp

https://www.england.nhs.uk/statistics/statistical-work-areas/covid-19-daily-deaths

https://www.gov.uk/government/publications/covid-19-track-coronavirus-cases.

## Acknowledgments

We would like to thank Anne Baker from the Office for National Statistics for very kindly providing her input to the COVID-19 related fatality data. We appreciate the work of J.D. Smith, B.M. Barratt, G.W. Fuller and team that captured the levels of PM2.5 exposure in London Underground.

## Author contributions

The authors confirm contribution to the paper as follows: study conception and design: M Sasidharan, A. Singh, M. Eskandari Torbaghan, A. K. Parlikad; analysis and interpretation of results: M Sasidharan, A. Singh; draft manuscript preparation: M Sasidharan, A. Singh, M. Eskandari Torbaghan. All authors reviewed the results and approved the final version of the manuscript.

## Data and materials availability

The data collected for this analysis are all publicly accessible and is available in the main text or the supplementary materials

## Supplementary Materials

Materials and Methods

Figures S1-S2

Table S1

